# Development and Validation of a Machine Learning Model That Uses Voice to Predict Aspiration Risk

**DOI:** 10.1101/2025.05.04.25326112

**Authors:** Cyril Varghese, Jianwei Zhang, Sara A. Charney, Abdelmohaymin Abdalla, Elizabeth Reeves, Stacy Holyfield, Adam Brown, Michelle Higgins, Hunter Stearns, Julie Liss, Nan Zhang, Diana Orbelo, Rebecca Pittelko, Lindsay Rigelman, Victor E. Ortega, David G. Lott, Visar Berisha

## Abstract

**Background:** Aspiration causes or aggravates a variety of respiratory diseases. Subjective bedside evaluations of aspiration are limited by poor inter-and intra-rater reliability, while gold standard diagnostic tests for aspiration, such as video fluoroscopic swallow study (VFSS) and fiberoptic endoscopic evaluation of swallowing (FEES), are cumbersome or invasive and healthcare resource intensive.

**Objective:** To develop and validate a novel machine learning algorithm that can analyze simple vowel phonations, to aid in predicting aspiration risk.

**Methods:** Recorded [i] phonations during routine nasal endoscopy from 163 unique patients were retrospectively analyzed for acoustic features including pitch, jitter, shimmer, harmonic to noise ratio (HNR), and others. Supervised machine learning (ML) was performed on the vowel phonations of those at high-risk for aspiration versus those at low-risk for aspiration. Ground truth of aspiration risk classification for model development was established using VFSS. The performance of the ML model was tested on an independent, external cohort of patient voice samples. The performance of trained Speech Language Pathologists (SLPs) to categorize high versus low-risk aspirators by listening to phonations was compared against the ML model.

**Results:** Mean ML risk score for those with the ground truth of high versus low aspiration risk was 0.530± 0.310 vs 0.243±0.249, which was a significant difference (0.287, 95% CI: 0.192-0.381) p<0.001. In the development cohort, the model showed an area under the curve (AUC) for the Receiver Operator Characteristic (ROC) of 0.76 (0.67-0.84) with specificity of 0.76 and F1 score of 0.63. The performance of the model in an external testing cohort was comparable, with AUC of 0.70 (0.52-0.88) with a specificity of 0.81, and F1 score of 0.67. The ML model had better accuracy, sensitivity, specificity, negative and positive predictive values compared to trained SLPs in classifying aspiration risk by evaluating vowel phonations.

**Conclusion:** High risk aspirators have quantifiable voice characteristics that significantly differ from those who are at a low risk for aspiration, as detected by a ML model trained to analyze sustained phonation and tested on an independent cohort. The ML model’s performance was superior to human experts in predicting aspiration risk by evaluating vowel phonations.

## INTRODUCTION

Abnormalities of the oropharynx or larynx can cause clinically relevant aspiration of oropharyngeal or gastrointestinal (GI) contents into the lungs [1]. Such abnormalities include tumor bulk, resection or radiation injury, pathological or age-related deterioration of nerves or muscles of the upper airways, esophageal abnormalities, and altered sensorium. Acute aspiration can cause significant injuries leading to pneumonias and Acute Respiratory Distress Syndrome (ARDS) [2, 3]. Chronic aspiration of GI contents can aggravate airway and parenchymal lung diseases such as chronic lipoid pneumonia, bronchiectasis, obliterative bronchiolitis, refractory asthma and pulmonary fibrosis [4], and is a major risk factor for transplanted lung rejection [5, 6].

Aspiration is common but is often undetected and usually only suspected after significant pulmonary damage has occurred. Despite its high prevalence in ambulatory and hospitalized patients [7–9], proactively detecting aspiration risk to prevent downstream sequelae is challenging. The most widely used hospital-based screening method to rule out anterograde aspiration (aspiration that occurs during swallowing) are nursing or speech-language pathologist (**SLP**) administered bedside swallowing evaluations (**BSEs**). BSEs vary across institutions but often include listening for a wet cough or wet voice quality after patients swallow varying amounts of liquid. Due to inter-rater/intra-rater reliability issues inherent with these subjective assessments [10], the sensitivity for BSEs ranges from 27% to 85%, with specificities ranging from 50-80% [11], and poor predictive values [12]. BSE results inform referral decisions for gold standard confirmatory testing including a videofluoroscopic swallow study (**VFSS**)[13] and fiberoptic endoscopic evaluation of swallowing (**FEES**)[14]. The VFSS exposes patients to radiation, requires coordinated participation, and can be challenging if patients are acutely ill, delirious, or have mobility issues, ex: in Intensive Care Units (**ICU**s). FEES is an invasive and uncomfortable evaluation where a scope is inserted through the nose and oropharynx to visualize the larynx during swallowing. Both VFSS and FEES are resource intensive, requiring specialized equipment and the expertise of radiologists, laryngologists, and SLPs for administration and proper interpretation, limiting wide scale deployment as screening tests.

Consequently, there is an *unmet need* for an objective bedside screening test that is easily administered and provides a valid indication of aspiration risk to support decisions regarding referral for VFSS/FEES.

Aspiration involves contact of GI/oropharyngeal contents with the vocal folds as they move through the airway and into the lungs. While perceptual-acoustic studies of postprandial phonation have shown some limited evidence of acute aspiration immediately after a swallow [15–17], chronic exposure to GI/oropharyngeal contents is likely to degrade the mucosal surfaces in ways that manifest as changes to the vocal folds’ vibratory and acoustic characteristics. Exposure to GI contents can induce histopathological changes to the vibratory margins of the vocal folds leading to changes in vocal quality[18]. We therefore anticipate that aspirators will exhibit changes in vocal quality relative to non-aspirators either due to underlying pathophysiology or as a consequence of chronic aspiration. In this study, we evaluated voice samples available from Mayo Clinic’s Otolaryngology clinical practices to develop, validate and externally test an ML algorithm that analyzes voice to predict aspiration risk. *<u>We hypothesize that increased risk of aspiration may be associated with changes in human voice quality that can be detected by machine learning (ML) approaches for the objective detection of aspiration risk</u>*.

## METHODS

The workflow for voice data curation, ML model development and validation is shown in **Fig 1**.

**Figure 1:**
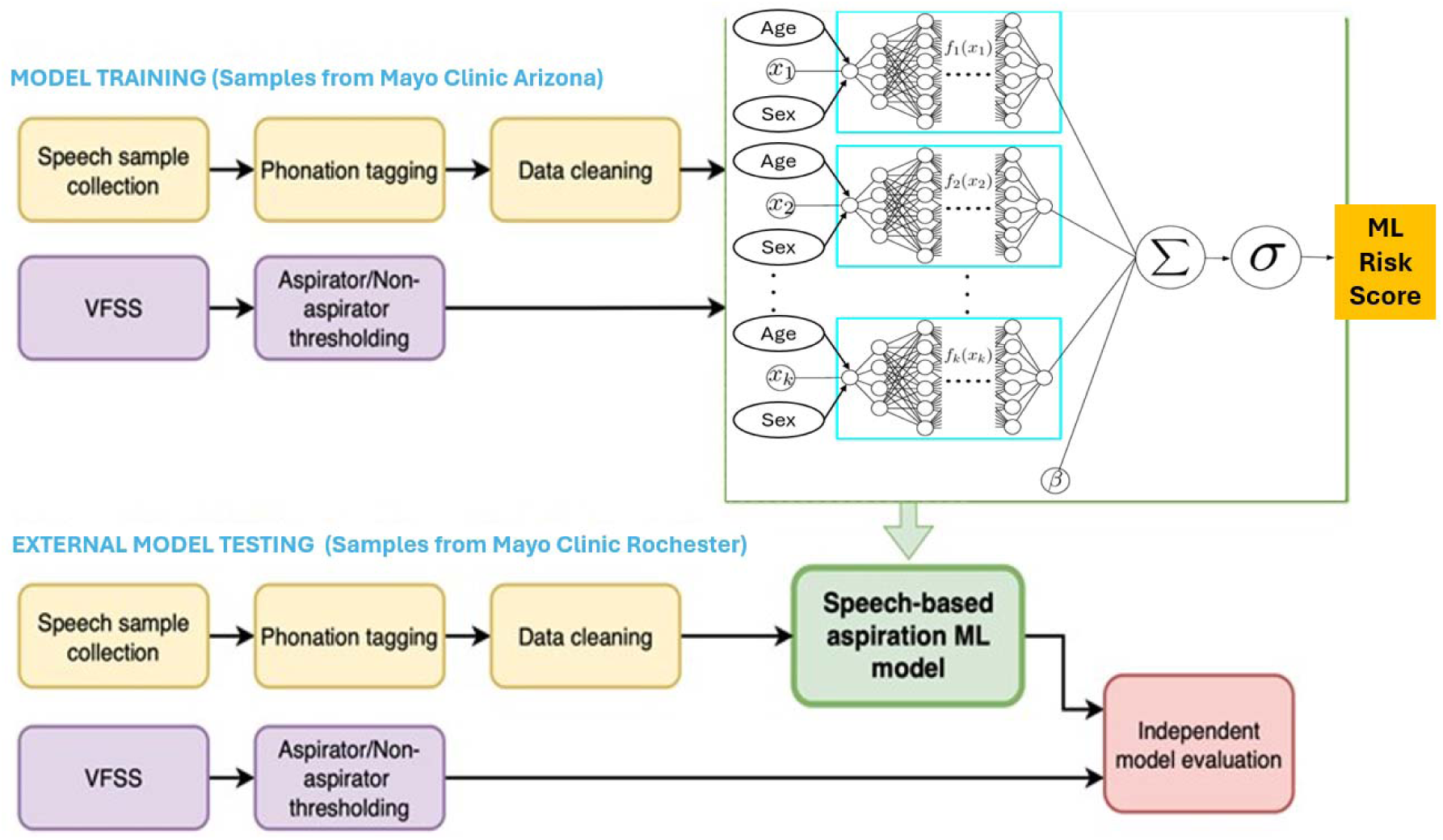
A graphical representation summarizing the workflow for this study from voice sample collection, phonation tagging, and subsequent ML model development based on gold standard testing (VFSS determined aspiration). Subsequently, independent model evaluation was performed wherein aspiration risk category based on VFSS was unknown to the model. Contained within the green box is a schematic of the neural additive model (NAM) [23] trained to estimate risk of aspiration utilizing 33 voice features, in full cohort (N=163), while adjusting for age and sex, the main confounders between the groups.⍰: summation of all sub-feature network outputs and learnable offset β; σ:represent the sigmoid function 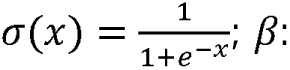 learnable offset of NAM model.

## ETHICS APPROVAL DECLARATION

This study was performed in accordance with the Declaration of Helsinki. This human study was approved by Mayo Clinic IRB.

### Voice Data Collection

Voice recordings collected during laryngoscopyic exams at Mayo Clinic Arizona (MCA), Otolaryngology clinics were extracted. All voice samples had been originally recorded during routine clinical workflow, using the lapel microphone (of the Pentax Laryngeal Strobe system) clipped near the clavicle. Endoscopy and VFSS databases from 1/2020 through 7/2021 were curated to identify patients with VFSS, an endoscopy exam, and voice recordings to identify 332 consecutive patients with both VFSS and voice recordings that were within 1 year of each other. Notably, since this is a retrospective study, voice sample collections were not specifically protocoled as would be done in a prospective study. Three SLPs (15 years combined experience) screened and manually excluded (Audacity 3.1.3) recordings with background noise, overlapping sounds (e.g., multiple speakers, machine sounds), or low volume. The samples were discarded if SLPs felt they were unable to reliably tag and label the individual samples, per the protocol below. Robust recordings were analyzed without computerized pre-processing to avoid artifacts from being introduced into the signal analysis pipeline. These samples were exported as.wav files and PRAAT (version 6.2.15) was used to tag the samples with specific labels, including sustained vowel phonation, cued speech, reading, and spontaneous speech. To maximize the number of participants used for training, o*nly [i] vowel phonations were analyzed*, as these were most consistently collected across all patients, during routine clinical practice. Focusing on a single vowel type also reduced variability arising from differences in speech elicitation. Reducing variability ensures that observed differences in acoustic features reflect true underlying vocal characteristics rather than differences in speaking style or task conditions. Of 772 voice recordings from 332 patients, 283 recordings (of multiple sustained [i] phonations) from 163 patients were analyzed. Three clips of the [i] vowel phonations lasting a minimum of 0.5 seconds from the central portion of each recorded clip per patient were used to develop the ML model.

### Clinical Data Collection

Clinical data extracted included aspiration risk factors, such as upper airway involvement (vocal fold disease, head and/or neck surgery, radiation exposures), esophageal diseases (strictures, impaired motility, GERD), neurological compromise (stroke, neuromuscular, neurodegenerative or peripheral nerve disease), body mass index (BMI), and obstructive sleep apnea.

SLPs evaluated VFSS results in training and validation cohorts with the Rosenbek Penetration-Aspiration Scale [19], (PAS) to classify patients as high-risk for aspiration (scores of 6-8, materials into the lower airways); moderate-risk (3-5 (materials in laryngeal vestibule and/or on vocal folds without being ejecting); and low-risk (1-2, no penetration or ejected out of airway). Notably, sustained /i/ phonations were recorded *separately from the VFSS*, on different days. As such the [i] phonations performed by patients were not influenced by simultaneous VFSS testing.

### Machine Learning and Statistical Analysis

In our initial model development and training, traditional voice features including pitch, jitter, shimmer, harmonics-to-noise ratio (HNR), cepstral peak prominence (CPP), and relative average perturbation (RAP) were extracted from tagged /i/ samples using audio processing toolboxes PRAAT (GNU General Public License Version 3, 29 June 2007) and Collaborative Voice Analysis Repository for Speech Technologies (COVAREP)[20]. For traditional voice features, parametric *t*-tests were used to compare those with high-and low-risk of aspiration. Comparisons of demographics and clinical characteristics between the groups were performed with Fishers’ exact test for categorical variables and Kruskal-Wallis rank sum test for continuous variables. These analyses were performed with the arsenal package (R4.2.2 [R Foundation for Statistical Computing, Vienna, Austria]). 0.05 was chosen as the cut-off criterion for statistical significance.

Neural additive models (NAM) [21]were used to classify the voice data and analyze the [i] phonation data of 163 patients. The following dependencies were used for ML coding in Python3: PyTorch, numPy and sciPy. For NAM, we included 33 features that are well described to have validity in characterizing pathological voice states [22–24]. Five-fold cross-validation and Recursive Feature Addition (RFA) [25, 26] was employed to identify the features required to effectively distinguish high-risk from low-risk aspirators and to estimate model performance.

### Independent Model Testing

The development NAM model was trained on voice data from Mayo Clinic in Arizona (MCA), while independent validation of the model was performed with voice data of patients from another distinct Mayo Clinic site: Mayo Clinic in Rochester (MCR). The clinicians, recordings, equipment/rooms, geographic location, and patient demographics of the external testing cohort (MCR) were independent of the training cohort (MCA). This allowed us to evaluate the out-of-distribution generalizability of the model, ensuring it’s focus on aspiration and not site-specific confounders. The selection, extraction, tagging, and processing of these externally collected voice samples used the *same* methodology as described previously for the development cohort. ML risk score for each patient in the testing cohort was calculated in a blinded fashion by the model by analyzing [i] phonation clips in the context of the patient’s age and sex. The gold standard designation (i.e., aspiration risk category based on VFSS) or clinical information was not known to the investigators running the ML code. Statistical analyses including sensitivity, specificity, receiver operator characteristics (**ROC**) were calculated to assess the model’s performance on both the testing and development cohorts.

### Comparing Human Expert Raters with ML Model

Four SLPs, from the two medical centers (40 years combined experience), blinded to medical history and aspiration risk, classified patients as at high-or low-risk for aspiration based on perceptual judgements of the [i] phonation clips. This is identical to the information that was provided to the ML model in testing. Notably, the moderate aspiration risk group is not included in the test set, as the goal of testing is to observe if this model developed on retrospective voice data can discriminate between high and low risk aspirators. The inter-rater reliability for the human raters were assessed using pairwise Cohen’s kappa. The sensitivity, specificity, positive/negative predictive values and accuracy of the human raters and the ML model for predicting high versus low-risk aspirators were calculated.

## RESULTS

### Feature Analysis

We initially analyzed retrospective samples from 87 patients (19 high-risk aspirators, 10 medium-risk aspirators, and 58 low-risk aspirators) with an average of approximately 2 voice files per patient. Non-parametric analysis of variance of demographic and clinically relevant risks of aspiration in the pilot data revealed that only age and sex were different among the groups. On age and sex-matching prior to analysis, 17 high-risk aspirators were found to be age-and sex-matched to 17 low-risk aspirators in our exploratory cohort. Of the traditional voice features that were explored, high-risk aspirators were found to have higher jitter and shimmer but lower harmonics richness factor than low-risk aspirators (**Figure 2**).

**Figure 2:**
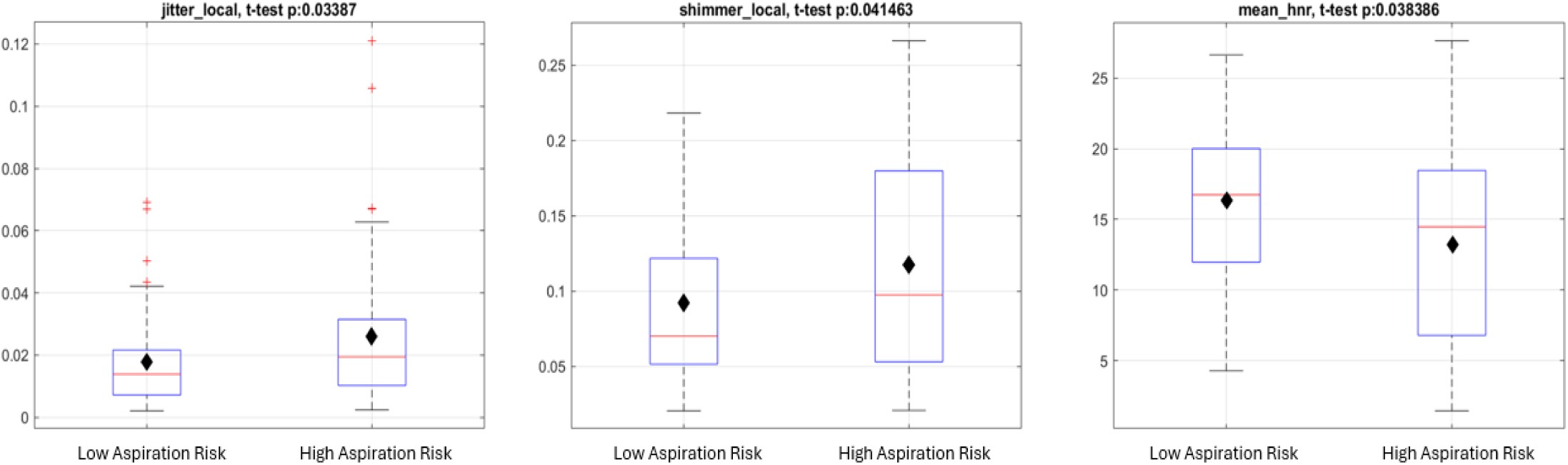
Pilot data of 17 age/sex matched aspirators with 17 non-aspirators, reveal that aspirators had a higher mean jitter (mean difference 0.0085, 95% CI: 0.000756-0.0161), shimmer (mean difference 0.0266, 95% CI: 0.00228-0.0506) and lower harmonics-to-noise ratio (HNR) (mean difference 3.1, 95% CI: 0.603-5.6). Means represented by black diamonds. Shimmer is the cycle-to-cycle variability in amplitude while jitter is the cycle-to-cycle variability in frequency.

### Development of ML Model and Internal Cross-Validation

Our primary ML model development and training cohort evaluated 163 patients within the three groups (47 high-risk aspirators, 17 moderate-risk aspirators, and 99 low-risk aspirators) that significantly differed by sex, age, and BMI (with high-risk aspirators having a lower BMI than low-risk aspirators). Additional differences included more reported solid and liquid dysphagia symptoms and a higher frequency of structural changes to head and neck anatomy in the high-risk group than in the low-risk group, as expected in an Otolaryngology population (**Table 1**).

**Table 1:** Baseline characteristics of training cohort.

|  | **High**<br>**Aspiration**<br>**Risk (N=47)** | **Low**<br>**Aspiration**<br>**Risk (N=99)** | **Moderate**<br>**Aspiration**<br>**Risk (N=17)** | **p value** |
| --- | --- | --- | --- | --- |
| **Sex** | | | | $< 0.001^1$ |
| Male | 39 (83.0%) | 45 (45.5%) | 9 (52.9%) |  |
| Female | 8 (17.0%) | 54 (54.5%) | 8 (47.1%) |  |
| **Age at Scope Exam** | | | | $< 0.001^2$ |
| Mean (SD) | 72.3 (9.9) | 62.9 (12.6) | 75.1 (9.0) |  |
| Median | 73.0 | 61.0 | 77.0 |  |
| Q1, Q3 | 67.0, 78.0 | 55.0, 72.0 | 65.0, 82.0 |  |
| Range | 40.0 - 88.0 | 31.0 - 90.0 | 62.0 - 89.0 |  |
| **BMI** | | | | $0.009^2$ |
| Mean (SD) | 24.1 (3.2) | 27.0 (5.3) | 26.4 (7.7) |  |
| Median | 23.8 | 26.8 | 25.3 |  |
| Q1, Q3 | 22.0, 26.1 | 23.4, 29.7 | 21.3, 29.4 |  |
| Range | 18.7 - 31.2 | 17.0 - 44.2 | 16.0 - 48.9 |  |
| **Dysphagia** | | | | $0.059^1$ |
| No | 4 (8.5%) | 25 (25.25%) | 2 (11.8%) |  |
| Yes | 40 (85.1%) | 72 (72.7%) | 14 (82.4%) |  |
| NA | 3<br>(6.4%) | 2 (2%) | 1 (5.9%) |  |
| **Esophageal disease group** |  |  |  | 0.551<sup>1</sup> |
| Clinical GERD | 23 (48.9%) | 46 (46.5%) | 5 (29.4%) |  |
| Other esophageal disease | 3 (6.4%) | 6 (6.1%) | 0 (0.0%) |  |
| Multiple esophageal diseases | 2 (4.3%) | 3 (3.0%) | 1 (5.9%) |  |
| GERD on impedance/manometry studies | 1 (2.1%) | 0 (0.0%) | 0 (0.0%) |  |
| No esophageal disease | 9 (19.1%) | 24 (24.2%) | 4 (23.5%) |  |
| NA | 9 (19.1%) | 20 (20.2%) | 7 (41.2%) |  |
| **OSA** |  |  |  | 0.871<sup>1</sup> |
| Compliant with CPAP | 8 (17.0%) | 19 (19.2%) | 1 (5.9%) |  |
| Not compliant with CPAP | 2 (4.3%) | 5 (5.1%) | 1 (5.9%) |  |
| Compliance not reported | 2 (4.3%) | 4 (4.0%) | 0 (0.0%) |  |
| No OSA | 35 (74.5%) | 71 (71.7%) | 15 (88.2%) |  |
| **Neurological illness** |  |  |  | 0.135<sup>1</sup> |
| CVA without any deficit | 1 (2.1%) | 4 (4.0%) | 0 (0.0%) |  |
| ALS | 0 (0.0%) | 1 (1.0%) | 0 (0.0%) |  |
| Neuromuscular diseases | 3 (6.4%) | 0 (0.0%) | 0 (0.0%) |  |
| CVA with dysphagia only | 1 (2.1%) | 0 (0.0%) | 0 (0.0%) |  |
| CVA with residual neuro deficit | 1 (2.1%) | 0 (0.0%) | 0 (0.0%) |  |
| None | 41 (87.2%) | 94 (94.9%) | 17 (100%) |  |
| **Vocal fold disease** |  |  |  | 0.338<sup>1</sup> |
| Yes | 23 (48.9%) | 35 (35.4%) | 8 (47.1%) |  |
| No | 22 (46.8%) | 62 (62.6%) | 9 (52.9%) |  |
| NA | 2 (4.3%) | 2 (2.0%) | 0 (0.0%) |  |
| **Head and neck anatomical disease** |  |  |  | 0.013<sup>1</sup> |
| Surgery | 6 (12.8%) | 13 (13.1%) | 2 (11.8%) |  |
| Cancer | 1 (2.1%) | 1 (1.0%) | 0 (0.0%) |  |
| Radiation | 0 (0.0%) | 2 (2%) | 0 (0.0%) |  |
| Multiple head and neck anatomical diseases | 25 (53.2%) | 22 (22.2%) | 5 (29.4%) |  |
| None | 15 (31.9%) | 61 (61.6%) | 10 (58.8%) |  |

1. Fisher's Exact Test for Count Data 2. Kruskal-Wallis rank sum test - NA: Not reported by patient or not found in chart review

Baseline characteristics for the age-and sex-matched cohort of 36 high-risk aspirators and 36 low-risk aspirators (N=72) showed BMI and incidence of head and neck disease was different between the groups (**Table Supplemental 1**).

Because age and sex influence voice, and were different between the groups, they were controlled for in the NAM (**Figure 1**). The model utilized 33 extracted features to differentiate between aspirators and controls (**Table Supplemental 2**). Each sub-network within the model processed a single extracted voice feature along with the corresponding sample sex and age as inputs (**Figure 1**). The sub-network output was a single scalar reflecting the adjustments made for age and sex within the feature. The whole NAM output for aspiration risk was a scalar ranging from 0 to 1. High-and low-risk aspirators were clearly distinguished: high-risk aspirator score=0.530 ([SD]=0.310) versus low-risk aspirator risk score of 0.243 (SD=0.249); mean risk difference between groups was 0.287 (95% CI: 0.192-0.381), p<0.001 (**Figure 3A**). Moderate-risk aspirators had a risk score between high-risk aspirators and low-risk aspirators, although this difference when compared to the other groups was not significant. RFA showed an elbow point at 7 features (**Figure 3B**), revealing that the most significant voice features adding to the model’s discriminability were the average fundamental frequency and standard deviation (F0_mean and F0_std [standard deviation]), the maximum fundamental frequency during phonation (Max Pitch), the standard deviation of the quai-open quotient that measures the proportion of the glottal cycle when the glottis is open (QOQ_std), the average of the harmonic richness factor (HRF_mean), the average of the cepstral peak prominence (CPP_mean), and the standard deviation of the cepstral peak prominence (CPP_std).

**Figure 3:**
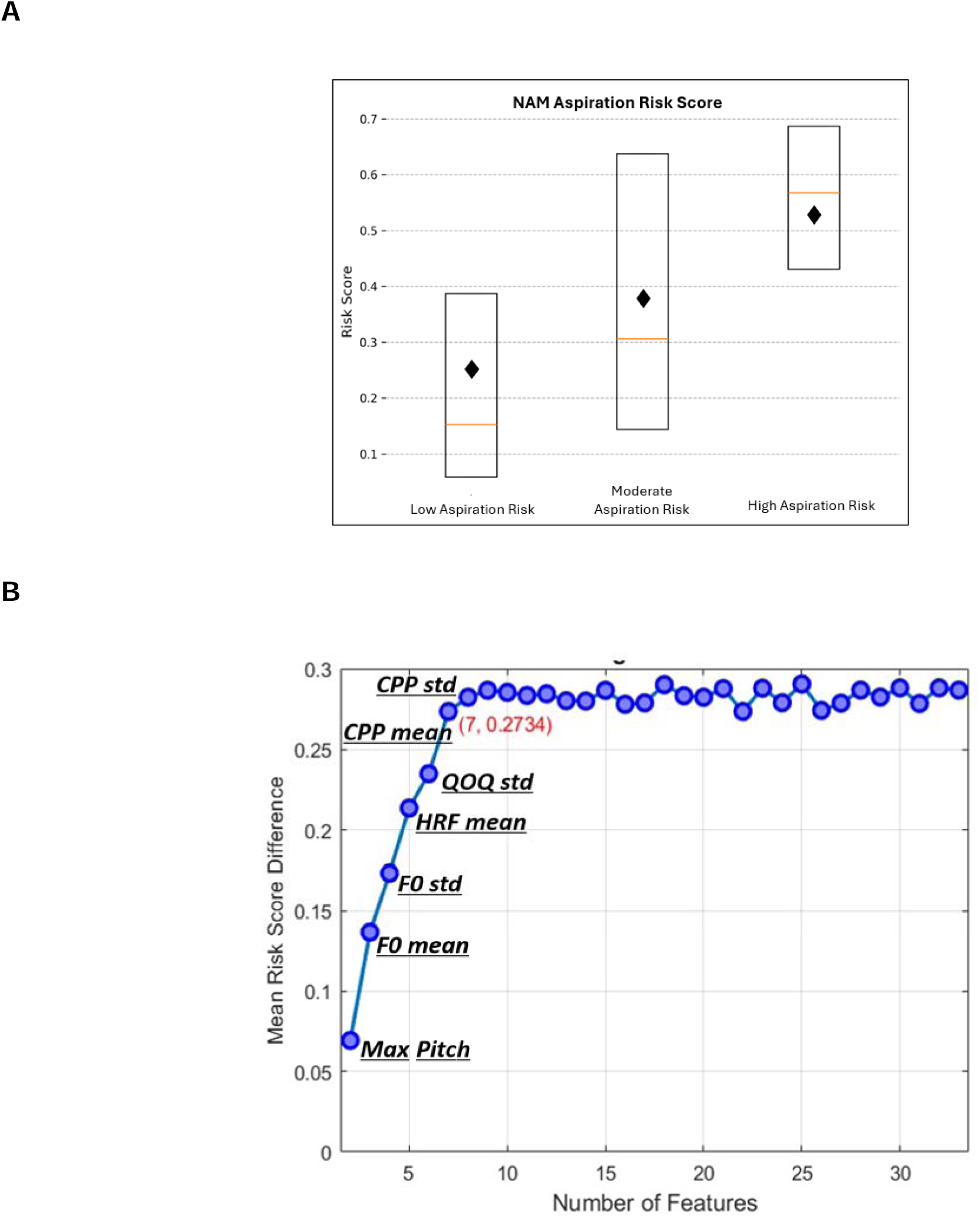
**A**: High-risk Aspirator Neural Additive Model (NAM) risk score 0.530 ± 0.31 (mean ± std) versus low-risk aspirator risk score of 0.243±0.249 (mean ± std); Mean risk difference between groups 0.287 (95% CI: 0.192-0.381), p<0.001. Means represented by black diamonds. Moderate-risk aspirators score fell between high and low risk aspirators. **B:** The Recursive Feature Addition (RFA) method was employed to identify the minimal set of features required to effectively distinguish between high and low-risk aspirators. This method initiates with a single feature and iteratively adds the voice feature that enhances the performance of the existing set. The key metric used to assess the performance of the feature set is the mean risk score difference.

A description of these features and the other 33 features analyzed in the NAM is in **Table Supplemental 2**.

### External Testing of the ML model

The external cohort contained 19 high-and 16 low-risk aspirators based on VFSS, which totaled <u>24% of the sample size</u> of high and low-risk aspirators in the development cohort. The demographics between the training and testing cohort were different. For example, the testing cohort had a higher proportion of female high-risk aspirators (20% vs 5.5%), and a lower proportion of male low-risk aspirators (8.6% vs 30.8%), p=0.005. The testing cohort was older (70.3±13.4 vs 65.9±12.6 years, p=0.029) The mean ML risk score output for the high-risk aspirators was significantly higher than the low-risk aspirators (Mean (SD):0.469 (0.327) vs 0.245 (0.265), p =0.047) (**Figure 4A**). The optimal ML risk score cut-off to distinguish high-from low-risk aspirators by analyzing the training cohort was 0.35 based on the Youden index (which is calculated as sensitivity+specificity-1 and determines the optimal cut-off for balancing sensitivity and specificity). Despite the demographic, the AUC for the ROC for the testing cohort was 0.697 (0.517-0.878) compared to AUC of 0.755 (0.666-0.843) for the training cohort (**Fig. 4B**). The precision, recall, F1 score and specificity of the model for the testing cohort were 0.79,0.57, 0.67 and 0.81 respectively **(Fig. 4C).**

**Figure 4:**
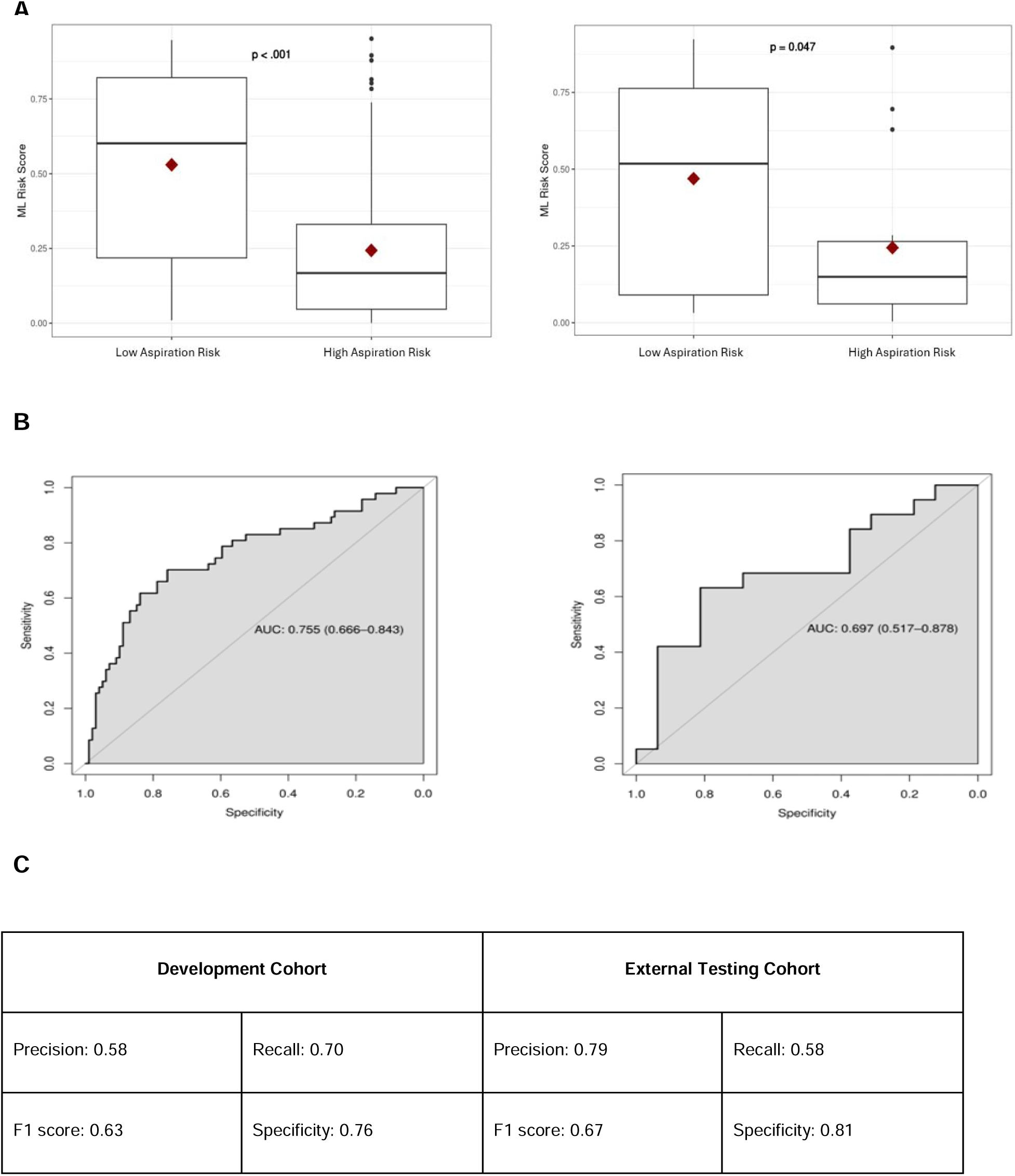
**A:** Box plot of ML Risk Score for training versus validation cohort. **B:** Receiver Operator Curve for development versus external testing cohort. AUC: Area under the curve. **C:** Analyses evaluating the performance of the model on training versus validation cohort. Where, F1 score= 2*Precision*Recall/ (Precision + Recall)

### Human Raters versus ML model

Other than two raters that had very good agreement (pairwise Cohen’s Kappa of 0.8) the rest had fair agreement (Choen’s Kappa range 0.34-0.59) (**Table 2 A**). In classifying patients as a high-or low-risk aspirator by analyzing [i] phonations, the ML model had higher accuracy than human raters’ range (69% vs 46-60%), sensitivity (58% vs 32-47%), PPV (79% vs 50-78%) and NPV (62% vs 41-54%) (**Table 2 B**). Other than one rater the ML model also had better specificity compared to human raters (81% vs 44-88%).

**Table 2:**
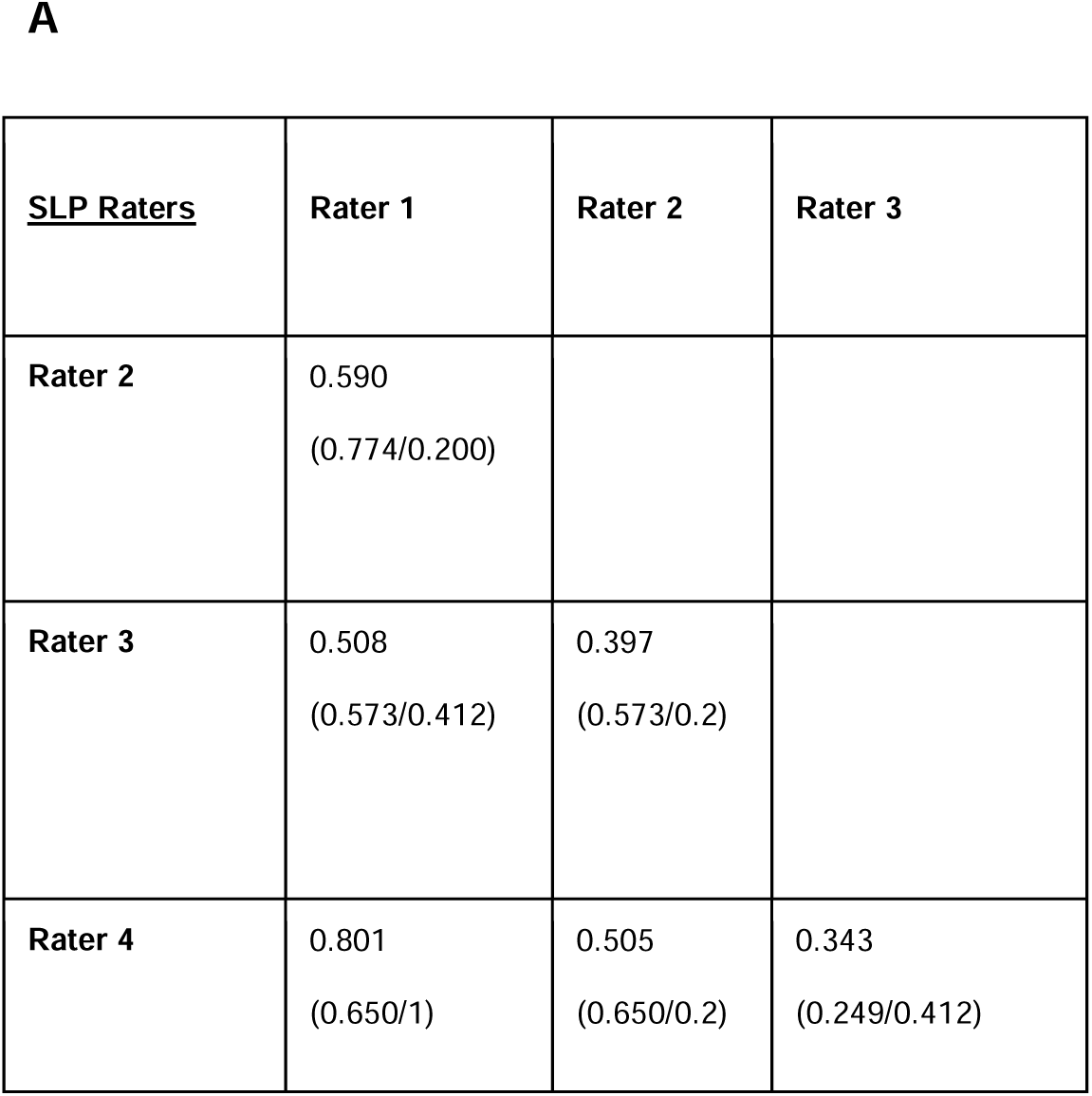
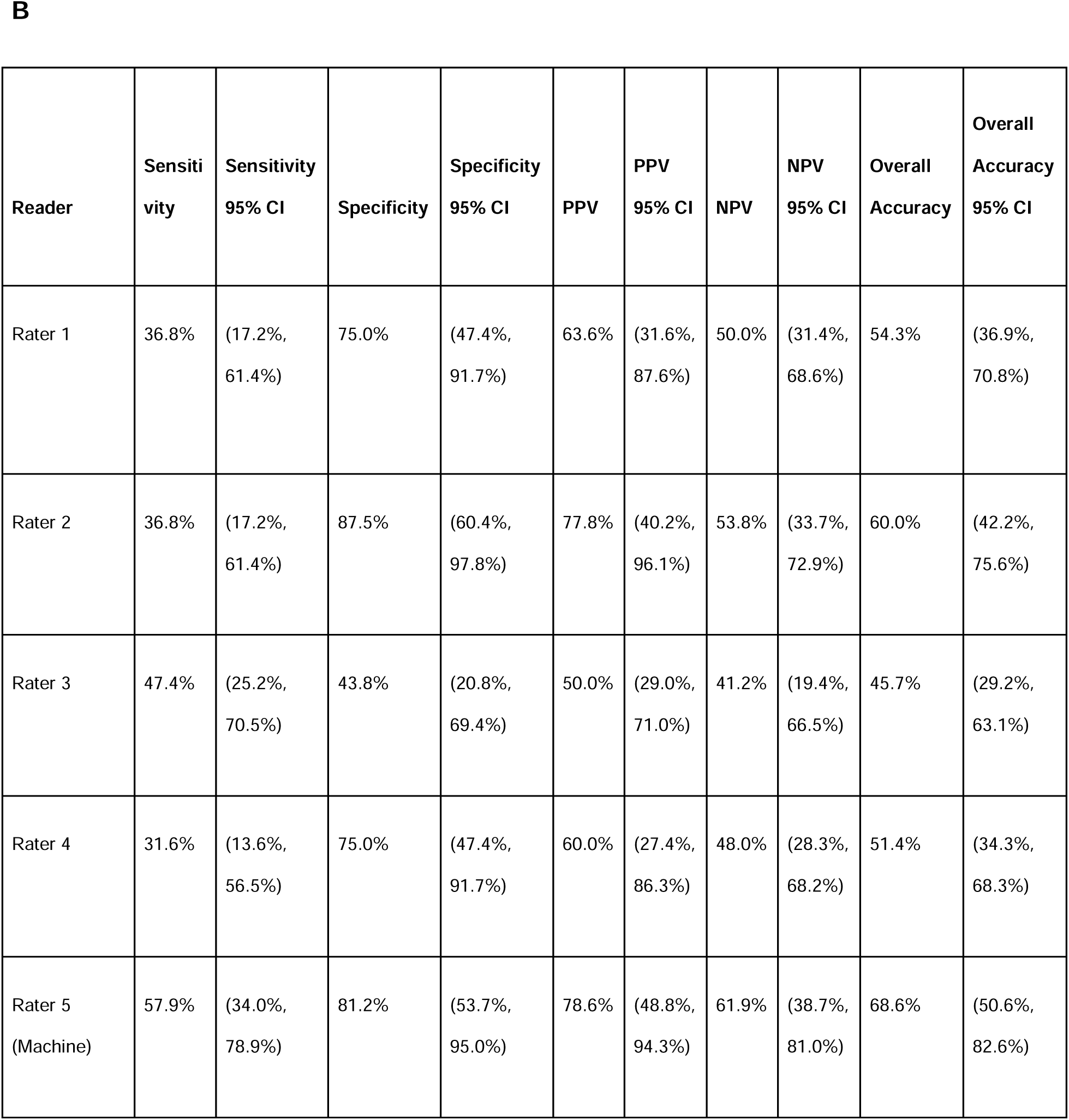
A:Inter-rater reliability pairwise Kappa coefficient between human SLP raters. In parenthesis inter-rater reliability between raters, as they made predictions of (high-risk/low risk) aspirators **B:** Blinded Human (raters 1-4) and Machine (rater 5) raters’ ability to predict aspiration risk by listening to phonations against the ground truth (based on VFSS).

## DISCUSSION

While acutely or chronically having ingested materials enter the larynx alters the quality of voice, human judgment of voice changes [27, 28] or even validated tools for perceptual assessments are unreliable screening methods to detect aspiration risk [29, 30]. There is a need for tools to objectively quantify aspiration-related voice changes in patients [15–17]. In a study of 93 patients, Relative Average Perturbation (RAP) combined with HNR increased sensitivity of detecting aspiration in the five voice features analyzed [15]. A similar study of 165 patients demonstrated that of 8 voice features evaluated, RAP was most distinguishing measure between aspirators and non-aspirators [16]. Recently, a study of 198 patients used ML to detect aspiration by analyzing postprandial voice [17]. However, all these studies were single center studies without an external testing cohort. The focus of these studies was on the effects of anterograde aspiration on voice as ingested materials contact the vocal folds in real time.

However, current bedside swallow evaluations also screen for aspiration by making the patient swallow. There are no available tests to objectively detect aspiration risk *without having the patient swallow liquids or solids*. This can be a limitation for frail, immobile, delirious patients who are already at a considerable risk for aspiration, as in the ICU. An objective screening test for aspiration risk that only requires simple phonation could be useful in these scenarios. Such a method could facilitate frequent longitudinal testing, refining referral for confirmatory testing with VFSS or FEES.

Unlike other studies in this space, we tested the performance of our model in an independent external testing cohort. Previous work has shown that external testing of models is critical as it can be difficult to train clinical speech models that generalize [31, 32] 33]. Our testing cohort was from a geographically distinct clinic where retrospective voice samples were collected from patients who were demographically different from our training cohort. Despite these differences, the model’s performance did not change significantly (**Figure 4**), suggesting that the model is not fragile. Fragile models that are over-fitted in development cohorts, tend to perform poorly in external cohorts, and therefore can seldom be used effectively in clinical practice. Finally, we explored if the ML tool added value as a preliminary test to predict aspiration risk by comparing its performance to SLPs. We found that the inter-rater reliability among our SLP raters was only “fair”, similar to reports in literature for other perceptual evaluations.

There is an inherent tradeoff between purely data-driven model development and approaches that incorporate domain expertise. While deep learning models typically require very large datasets to automatically learn relevant features, simpler supervised machine learning methods can achieve strong performance with smaller sample sizes when domain experts guide the feature selection and validation process [34]. One of the benefits of NAM used in this evaluation is the sequential way with which features can be considered and confounding variables controlled, thereby improving explainability of the model [21]. While simpler models like logistic regression can classify by accounting for basic covariates such as age and sex, they are limited in capturing non-linear relationships and feature-specific interactions. The NAM’s feature-specific networks capture complex, non-linear interactions while controlling for relevant patient characteristics.

This study has several limitations. The relationship between voice and aspiration is likely a complex bidirectional relationship that cannot be fully elucidated with a retrospective analysis. Our population of ENT patients have other reasons related or unrelated to aspiration for their voice to be altered (e.g., radiation-induced vocal fold scarring). We controlled for age and sex (predictors known to covary with voice) using our neural additive model. However, other clinical factors like BMI and anatomical head/neck disease are known to covary with aspiration (the response variable) either as a cause or an effect and therefore cannot be easily controlled by generalized regression models in a retrospective analysis. Also, we also focused on anterograde aspiration as confirmed by VFSS. It is possible in our cohorts that participants experienced retrograde aspiration, wherein micro-aspiration of gastrointestinal contents occurs typically during sleep in the context of gastroesophageal reflux disease [4]. Because it can occur in the absence of any structural or functional abnormalities, VFSS/FEES are not sensitive diagnostics. This may have resulted in false positive high-risk errors in both SLP and ML classifications. The inclusion of indirect (reflux symptoms or esophageal impedance testing) and direct indicators of retrograde aspiration (e.g. pepsin and bile in bronchoalveolar lavage) in our model may further refine model performance beyond the AUC of.70 range. Additional fine-tuning may be achieved by including measures from connected speech and increasing diversity of clinical settings. While constrained by the retrospective voice data, this formative study motivates a prospective multi-site voice collection trial to better characterize voice and its relationship to aspiration.

The elbow point in the cross-validation analysis (**Figure 3B**) was attained at 7 features (relevant features highlighted in **Table Supplemental 2**). These features and interactions were the most predictive of aspiration risk, but only in the specific sample we evaluated, namely ENT patients. We expect that as we expand our databases to include more diverse patient samples from different clinical settings, other voice features may emerge as significant predictors. Our categorization of “high” and “low” aspiration risk was based on VFSS at one point in time. It could be claimed that this is not reflective of the patient’s true longitudinal aspiration status. This is why we were careful to label the categories in terms of “risk” rather than absolute diagnoses. Those categorized as high aspiration “risk” were widely separated from the low aspiration “risk” (PAS 6-8 vs PAS 1-2), and crossovers between these categories in our ENT practice although possible is unlikely. While our model had a superior performance compared to our expert SLP raters, it should be noted that SLPs do not make decisions about aspiration risk based on sustained phonations alone. However, this study highlights the potential benefit of ML to complement informed clinical decision making by experts. Finally, we do not claim that this ML tool can be used as a confirmatory diagnostic test like VFSS or FEES. Rather the goal of this study was to develop and validate a tool that can potentially serve as an easily deployable screening test used by bedside nurses and SLPs to more objectively screen for anterograde aspiration risk.

This study is the first of its kind to utilize ML techniques to quantify the quality of voice to estimate aspiration risk *without performing a simultaneous swallow evaluation*. We expect that ongoing development of such technology will facilitate the early identification of aspiration in a variety of clinical settings including ICUs, hospital wards, ambulatory clinics, and remote monitoring, which could potentially impact the mitigation and management of several acute and chronic pulmonary diseases.

## Abbreviations

SLPs: Speech Language Pathologists
ENT: Ear Nose & Throat or Otolaryngology
ML: Machine Learning
HNR: harmonic-to-noise ratio
RAP: relative average perturbation
VFSS: Video Fluoroscopic Swallow Study
PAS: Penetration Aspiration Scale
FEES: Fiberoptic Endoscopic Evaluation of Swallowing
ICU: Intensive Care Units
NAM: Neural Additive Model

## Data Availability

All data produced in the present work are contained in the manuscript

