## Supplemental Tables 1 and 2 for "Development and Validation of a Machine Learning Model That Uses Voice to Predict Aspiration Risk"

**Table Supplemental 1:** Baseline characteristics of training cohort when high-risk and low-risk aspirators were age and sex-matched

|  | **High Aspiration Risk (N=36)** | **Low Aspiration Risk (N=36)** | **p value** |
| --- | --- | --- | --- |
| **Sex** |  |  | 1.000^1^ |
| Male | 28 (77.8%) | 28 (77.8%) |  |
| Female | 8 (22.2%) | 8 (22.2%) |  |
| **Age at Scope Exam** |  |  | 0.636^2^ |
| Mean (SD) | 70.9 (10.2) | 70.1 (10.0) |  |
| Median | 73.0 | 71.0 |  |
| Q1, Q3 | 65.8, 77.2 | 66.8, 77.0 |  |
| Range | 40.0 - 88.0 | 40.0 - 90.0 |  |
| **BMI** |  |  | 0.003^2^ |
| Mean (SD) | 24.1 (3.0) | 27.6 (5.5) |  |
| Median | 23.7 | 27.0 |  |
| Q1, Q3 | 22.8, 26.0 | 23.8, 30.2 |  |
| Range | 18.7 - 31.2 | 17.0 - 44.2 |  |
| **Dysphagia** |  |  | 0.045^1^ |
| No | 3 (8.3%) | 9 (25.0%) |  |
| Yes | 30 (83.3%) | 27 (75.0%) |  |
| NA | 3 (8.3%) |  |  |
| **Esophageal disease group** |  |  | 0.888^1^ |
| Clinical GERD | 16 (44.4%) | 14 (38.9%) |  |
| Other esophageal disease | 3 (8.3%) | 3 (8.3%) |  |
| Multiple esophageal diseases | 1 (2.8%) | 1 (2.8%) |  |
| GERD on impedance/manometry studies | 1 (2.8%) | 0 (0.0%) |  |
| No esophageal disease | 7 (19.4%) | 11 (30.6%) |  |
| Not reported | 8 (22.2%) | 7 (19.4%) |  |
| **OSA** |  |  | 0.608^1^ |
| Compliant with CPAP | 6 (16.7%) | 10 (27.8%) |  |
| Not compliant with CPAP | 2 (5.6%) | 1 (2.8%) |  |
| Compliance not reported | 1 (2.8%) | 2 (5.6%) |  |
| No OSA | 27 (75.0%) | 23 (63.9%) |  |
| **Neurological illness** |  |  | 0.186^1^ |
| CVA without any deficit | 0 (0.0%) | 2 (5.6%) |  |
| Neuromuscular diseases | 2 (5.6%) | 0 (0.0%) |  |
| CVA with dysphagia only | 1 (2.8%) | 0 (0.0%) |  |
| neuro not mentioned | 3 (8.3%) | 1 (2.8%) |  |
| None | 30 (83.3%) | 33 (91.7%) |  |
| **Vocal fold disease** |  |  | 0.926^1^ |
| Yes | 17 (47.2%) | 15 (41.7%) |  |
| No | 17 (47.2%) | 19 (52.8%) |  |
| Not reported | 2 (5.5%) | 2 (5.6%) |  |
| **Head and neck anatomical disease** |  |  | 0.056^1^ |
| Surgery | 2 (5.6%) | 3 (8.3%) |  |
| Cancer | 1 (2.8%) | 0 (0.0%) |  |
| Radiation | 0 (0.0%) | 1 (2.8%) |  |
| Multiple head and neck anatomical diseases | 21 (58.3%) | 11 (30.6%) |  |
| None | 12 (33.3%) | 21 (58.3%) |  |

1.	Fisher’s Exact Test for Count Data

2.	Kruskal-Wallis rank sum test

NA:	Not reported by patient or not found on chart review

**Table Supplemental 2:** List of 33 voice features that were used for machine learning organized in clinically meaningful domains. The seven features that contributed most to the model’s discriminability are bolded.

| **Group** | **Feature** | **Description** |
| --- | --- | --- |
| Jitter Features    These measure variations in pitch and are indicators of vocal stability. Higher values of jitter measurements typically indicate greater variability, which is often associated with pathological voices. | jitter_local | Average absolute difference between consecutive periods, divided by the average period. |
|  | jitter_local_abs | Average absolute difference between consecutive periods, in seconds. |
|  | jitter_rap | Relative average perturbation, the average absolute difference between a period and the average of it and its two neighbors, divided by the average period. |
|  | jitter_ppq5 | Five-point Period Perturbation Quotient, the average absolute difference between a period and the average of it and its four closest neighbors, divided by the average period. |
|  | jitter_ddp | Average absolute difference between consecutive differences between consecutive periods, divided by the average period. |
| Shimmer Features    These measure variations in amplitude of the acoustic signal, reflecting vocal fold vibration regularity. Higher shimmer values suggest more amplitude variability, commonly found in voices with pathologies. | shimmer_local | Average absolute difference between the amplitudes of consecutive periods, divided by the average amplitude. |
|  | shimmer_local_db | Average absolute base-10 logarithm of the difference between the amplitudes of consecutive periods, multiplied by 20. |
|  | shimmer_apq3 | Three-point Amplitude Perturbation Quotient, the average absolute difference between the amplitude of a period and the average of the amplitudes of its neighbors, divided by the average amplitude. |
|  | shimmer_apq5 | Five-point Amplitude Perturbation Quotient, the average absolute difference between the amplitude of a period and the average of the amplitudes of it and its four closest neighbors, divided by the average amplitude. |
|  | shimmer_apq11 | 11-point Amplitude Perturbation Quotient, the average absolute difference between the amplitude of a period and the average of the amplitudes of it and its ten closest neighbors, divided by the average amplitude. |
|  | shimmer_dda | Average absolute difference between consecutive differences between the amplitudes of consecutive periods. |
| Pitch Features | **F0_mean** | **Average fundamental frequency, related to the perceived pitch of the voice.** |
|  | **F0_std** | **Standard deviation of the fundamental frequency, indicating how much pitch varies over time.** |
|  | min_pitch | Minimum fundamental frequency during phonation. |
|  | **max_pitch** | **Maximum fundamental frequency during phonation.** |
| Harmonic Features    These features reflect voice quality (clarity/richness) and can help identify abnormalities or pathologies. | hnr_mean | Mean harmonics-to-noise ratio, indicating the ratio of harmonic sound to noise in the voice signal. |
|  | hnr_std | Standard deviation of the HNR, reflecting variability in voice quality. |
|  | h1h2_mean | Mean difference in amplitude between the first and second harmonics. |
|  | h1h2_std | Standard deviation of the difference between the first and second harmonics. |
|  | **hrf_mean** | **Mean of the harmonic richness factor, a measure of how much energy is in the harmonics relative to the fundamental frequency.** |
|  | hrf_std | Standard deviation of the harmonic richness factor, reflecting variability in harmonic richness. |
| Glottal Source Analysis Features | peak_slope_mean | Average slope of the spectral peaks, indicative of how sharply energy dissipates |
|  | peak_slope_std | Standard deviation of the spectral peak slope, showing variability in how energy distribution changes over time. |
|  | mdq_mean | Average of the Maxima Dispersion Quotient (MDQ), often related to the rate of vocal fold closure. |
|  | mdq_std | Standard deviation of the MDQ, indicating the consistency of vocal fold closure. |
|  | naq_mean | Mean normalized amplitude quotient (NAQ), which relates to the efficiency of the glottal closure during phonation. |
|  | naq_std | Variability in NAQ, indicating how consistently the vocal folds are closing. |
|  | qoq_mean | Average quai-open quotient (QOQ), which measures the proportion of the glottal cycle during which the glottis is open. |
|  | **qoq_std** | **Standard deviation of the QOQ, indicating the consistency of glottal opening during phonation.** |
|  | psp_mean | Average peak slope of the spectrum, offering insight into how spectral energy is concentrated around the peak frequency. |
|  | psp_std | Variability in the spectral peak slope, which can reflect changes in vocal tract dynamics or resonance strategies. |
|  | **cpp_mean** | **Mean cepstral peak prominence, which is a measure of the prominence of the peak in the cepstral domain, indicative of voice quality.** |
|  | **cpp_std** | **Variability in CPP, indicating how consistently the voice can produce clear, strong cepstral peaks.** |
